# Phenotypes and Electrophysiological Features of Hereditary Neuropathy with Liability to Pressure Palsies (HNPP) According to Physical Activities: A Single Military Hospital Experience

**DOI:** 10.1101/2024.08.25.24312370

**Authors:** Eun Jin Kim

**Affiliations:** Department of Physical Medicine and Rehabilitation, The Armed Forces Capital Hospital, Seongnam-si, Gyeonggido, South Korea

**Keywords:** neurogenetics, neuropathy, peripheral neuropathology

## Abstract

**Background:** The typical symptom of HNPP is focal nerve palsy with complete or near complete recovery, and it often occurs during military service with upper extremity weakness due to a brachial plexus involvement. This study aimed to analyze the clinical features and neurophysiological examination results of HNPP occurring in the Korean military to determine whether the clinical presentations and electrodiagnostic patterns differ according to the trigger activity and to obtain useful information for the early diagnosis of HNPP.

**Methods:** Sixty-four patients with HNPP were included over an 11-year period, and medical records and electrophysiological tests were analyzed.

**Results:** Thirty-five patients presented with isolated compressive neuropathy and 18 patients presented with brachial plexopathy. Analysis of phenotypes according to triggering activity revealed that 13 of 16 patients with symptom onset after military training showed a type of isolated compressive neuropathy and that 11 of 20 patients with symptom onset after push-ups or after backpack carriages showed a type of brachial plexopathy. Six patients experienced symptoms following sleep, while four reported symptoms after trauma. Atypical features were observed in 11 cases (17.2%), including 3 lower lumbosacral radiculopathy-like presentation. Abnormal findings were observed in 91.2% of distal latency of the peroneal motor nerve.

**Conclusions:** The phenotype varied according to the physical activity that triggered HNPP onset in soldiers. These results are expected to help in the early diagnosis of HNPP in soldiers or those who develop it after physical activity.

**key messages:**

- This study underscores the importance of recognizing clinical manifestation of HNPP in military personnel, particularly those involved in physically demanding activities, to facilitate timely diagnosis and intervention.
- The phenotypes of HNPP in soldiers vary depending on the triggering activity, with isolated compressive neuropathy and brachial plexopathy being the most common presentations.
- Atypical clinical manifestations were observed, including presentations resembling Guillain–Barré syndrome and lumbosacral radiculopathy.
- Regardless of the location of symptoms, a peroneal motor conduction study can be valuable in diagnosing HNPP.
- These findings could significantly contribute to the early diagnosis of HNPP in soldiers.

## INTRODUCTION

Hereditary neuropathy with liability to pressure palsies (HNPP) is an autosomal dominant genetic disorder (PMP 22 gene) characterized by recurrent acute painless palsy, with the wrist, elbow, knee, and shoulder being the most susceptible areas.^1,2,3)^ Study on soldiers with HNPP has shown that the most common symptom is proximal upper extremity weakness due to brachial plexus injury, and brachial plexus involvement is relatively more common among soldiers compared to the reported 10%–30% occurrence in the general HNPP population. ^4,5)^ In particular, soldiers have been reported to develop brachial plexus injuries after training and subsequently be diagnosed with HNPP.^6)^ Previous studies reported that push-ups were the most common physical activity associated with brachial plexus injuries in military patients with HNPP. ^7,8)^

Interest in HNPP occurring in soldiers was focused on injury to the brachial plexus, but clinically, it shows various clinical features depending on the physical activity that causes symptoms. The study of the clinical features and electrophysiologic findings of HNPP is important for early diagnosis. Because the clinical presentation of HNPP varies depending on the nerve involved, there is a need to study the presenting phenotypes and electrodiagnostic patterns according to symptom-inducing activities. However, there is a lack of studies on the clinical presentation of HNPP in military personnel and the physical activities that trigger it.

This study aimed to analyze the phenotype of HNPP in soldiers according to the triggering activity and analyze the characteristics of the electrophysiological test to obtain information that will help diagnose HNPP early.

## METHODS

### Study subjects

This retrospective study included patients diagnosed with HNPP via electrophysiologic and genetic testing at the Armed Forces Capital Hospital in South Korea from 2012 to 2022. The diagnosis criteria for HNPP were limited to patients who met clinical and electrodiagnostic criteria^9)^ and had abnormalities confirmed by genetic testing.

### Data collection

The clinical characteristics, neurophysiological test, and genetic test results of the subjects were collected by searching medical records for 11 years.

Statistical analysis

The SPSS (version 11.0) statistical program was used for statistics, and the Student’s T-test was applied for statistical comparison of the left and right nerve conduction study results.

### Ethical consideration

The institutional review board of the Armed Forces Capital Hospital approved the study (AFCH 2023-05-001).

## RESULTS

Sixty-four patients were enrolled after checking their medical records, and the average age of the patients was 22.2 (18–44) years, with one of them being female. The ranks of the patients were 24 private second class soldiers, 19 private first class soldiers, 11 corporals, 5 sergeants, 4 noncommissioned officers (1 sergeant major, 2 master sergeants, and 1 sergeant first class), and 1 warrant officer.

The average height of patients was 174.7 (164–186) cm, the average weight was 70.0 (52–100) kg, and the average body mass index was 28.5 (15.4–55.6) kg/m^2^. The departments that the patients first visited included neurology (22 patients), orthopedic surgery (18 patients), rehabilitation medicine (15 patients), and neurosurgery (9 patients).

The timing of symptom onset in the patients was an average of 5 months prior to the initial consultation day, and five patients had symptoms prior to enlistment. There were 20 patients whose symptoms began within 1 month of enlistment, 17 whose symptoms began within 6 months of enlistment, and 6 whose symptoms began within 1 year of enlistment. Five patients had symptoms that started 1 year after enlistment, and 10 patients could not remember when the symptom started.

Fifty-four patients had symptoms in the upper extremity, 21 on the right, 23 on the left, and 9 bilaterally. The lower extremities were involved in nine patients, with five patients experiencing symptoms on the right side, two on the left side, and two bilaterally. Two patients had all four limbs involved. The most common initial symptom reported by patients was muscle weakness (38), followed by paresthesia (17), tingling (14), pain (10), and claw hand (2).

Twelve patients had a family history of acute nerve palsy. Twenty-nine patients had a history of nerve palsy, of whom two underwent surgical treatment for nerve compression (median nerve decompression and/or ulnar nerve decompression).

The patients were followed up for an average of 8.8 months, and 48 patients were discharged with mental and physical disabilities because they were unable to continue their military service due to physical problems. Table 1 summarizes the basic information and clinical presentation of the patients.

**Table 1.** Demographic and clinical findings of 64 patients with HNPP.

| Findings |  |
| --- | --- |
| Male/Female | 63 / 1 |
| Average age (years old) | 22.2 (18-44) |
| Rank |  |
| Sergeant first class/Master sergeant/Sergeant major/Warrant officer | 1/2/1/1 |
| Enlisted soldiers (PV1/PFC/Corporal/Sergeant) | 59 (24/19/11/5) |
| Physical condition |  |
| Average height (cm) | 174.7 (164-186) |
| Average weight (kg) | 70.0 (52-100) |
| Average BMI (kg/m<sup>2</sup>) | 28.5 (15.4-55.6) |
| Department of initial consultation |  |
| Neurology | 22 |
| Orthopedic Surgery | 18 |
| Rehabilitation Medicine | 15 |
| Neurosurgery | 9 |
| Average symptom duration (months) | 5 (0-72) |
| Symptom onset before enlistment | 5 |
| Symptom onset within 1 month of enlistment | 20 |
| From 1 month to 6 months after enlistment | 17 |
| From 6 months to 1 year after enlistment | 6 |
| Others | 15 |
| Location (Side) |  |
| Upper (Right/Left/Both) | 53 (21/23/9) |
| Lower (Right/Left/Both) | 9 (5/2/2) |
| Both Upper and Lower | 2 |
| Presenting manifestations |  |
| Muscle weakness | 38 (59.4%) |
| Sensory deficit | 17 (26.6%) |
| Paresthesia | 14 (21.9%) |
| Pain | 10 (15.6%)) |
| Claw hand | 2 (3.1%) |
| Family History of nerve palsy | 12 (18.8%) |
| Previous event of nerve compression | 29 (45.3%) |
| History of surgery for nerve compression | 2 (3.1%) |
| Discharge from military service due to physical disabilities | 48 (75.0%) |

### Triggering activities and phenotypes

The most common triggering activity for symptom onset was military training, including physical activities such as fitness training and basic military training such as shooting and obstacle course training, reported by 16 people. Thirteen patients reported symptom onset after push-ups, 7 after carrying backpacks, 6 upon waking up, 4 after trauma (shoulder dislocation, fracture, etc.), and 18 did not report any specific triggering activity.

The authors categorized patients into five phenotypes based on clinical presentation, referring to existing studies: ^2,3,10)^ 1) isolated compressive neuropathy, 2) brachial plexopathy, 3) polyneuropathy, 4) radiculopathy, and 5) multiple mononeuropathy.

The most common phenotype was isolated compressive neuropathy in 35 patients, followed by brachial plexopathy in 18, polyneuropathy in 5, radiculopathy in 4, and multiple mononeuropathy in 1. Among 35 patients with isolated compressive neuropathy, the ulnar nerve was most commonly involved in 14 patients, followed by the musculocutaneous nerve in 8, the radial nerve in 6, the peroneal nerve in 5, and the axillary and median nerves in 1 each.

The analysis of phenotypes according to the triggering activities showed the type of isolated compressive neuropathy in 13 out of 16 patients with symptom onset after military training (physical training and basic military training). The specific neural phenotypes of isolated compressive neuropathy included ulnar and musculocutaneous nerves in four patients each, radial and peroneal nerves in two patients each, and the axillary nerve in one patient. Of the remaining three, two had brachial plexus abnormalities and one had a type of radiculopathy.

In patients with symptom onset after push-ups or backpack carriages, the phenotype was most often brachial plexopathy (7 of 13 patients and 4 of 7 patients, respectively). Of the remaining six patients whose symptoms started after push-ups, two had a type of radial nerve, one had the musculocutaneous nerve, two had polyneuropathy, and one had radiculopathy. Of the remaining three patients whose symptoms started after backpack carriages, two had musculocutaneous nerve disorders and one had polyneuropathy.

In the six patients with symptom onset after sleep, radial nerve palsy and brachial plexus palsy were present in two patients each and musculocutaneous nerve palsy and peroneal nerve palsy were present in one patient each. In the 18 patients with symptom onset without any specific triggering activity, ulnar nerve involvement was the most common phenotype in 10 patients (Table 2).

**Table 2.** Triggering Activities and Presenting Phenotypes.

| Presenting Phenotype<br>N (%) | Triggering Activities<br>N (%) | Military<br>training | Push-ups | Backpack<br>carriages | Sleep | Trauma | Unknown |
| --- | --- | --- | --- | --- | --- | --- | --- |
|  |  | 16 (25.0) | 13 (20.3) | 7 (10.9) | 6 (9.4) | 4 (6.3) | 18 (28.1) |
| Isolated compressive neuropathy | 35 (54.7) | 13 | 3 | 2 | 4 | 1 | 12 |
| Ulnar nerve | 14 | 4 |  |  |  |  | 10 |
| Musculocutaneous nerve | 8 | 4 | 1 | 2 | 1 |  |  |
| Radial nerve | 6 | 2 | 2 |  | 2 |  |  |
| Peroneal nerve | 5 | 2 |  |  | 1 | 1 | 1 |
| Axillary nerve | 1 | 1 |  |  |  |  |  |
| Median nerve | 1 |  |  |  |  |  | 1 |
| Brachial plexopathy | 18 (28.1) | 2 | 7 | 4 | 2 | 1 | 2 |
| Polyneuropathy | 5 (7.8) |  | 2 | 1 |  | 1 | 1 |
| Radiculopathy | 4 (6.3) | 1 | 1 |  |  | 1 | 1 |
| Multiple mononeuropathy | 1 (1.6) |  |  |  |  |  | 1 |
| Sensory polyneuropathy | 1 (1.6) |  |  |  |  |  | 1 |

### Atypical clinical presentation

Three patients with a Guillain–Barre syndrome-like presentation were diagnosed with Guillain–Barre syndrome after presenting with chief complaints of decreased muscle strength or sensation in the upper extremities that occurred an average of 5.3 months before the hospital visit, with two of them treated with intravenous immunoglobulin (IVIG) without improvement.

Of the three patients who presented to the neurosurgery department with chief complaints of radiating pain to the lower extremities accompanied by lower back pain that occurred an average of 4.5 months before the hospital visit and showed a lumbosacral radiculopathy-like presentation, one patient had persistent pain after surgical treatment (partial hemilaminectomy and microdiscectomy) for herniated intervertebral disc

Three patients who presented with tingling sensations in the upper extremities that began on average 28.7 months before the hospital visit were found to have chronic sensory neuropathy. Two patients presented with chronic ulnar neuropathy, characterized by persistent symptoms lasting for more than 12 months.

### Electrodiagnostic study

After analyzing the nerve conduction studies of 64 patients diagnosed with HNPP, 47, 8, 7, and 2 were found to have sensorimotor polyneuropathy, sensorimotor polyneuropathy combined with brachial plexopathy, brachial plexopathy, and bilateral peroneal neuropathy, respectively.

When electromyography results were analyzed according to the triggering activities, 6/16, 5/13, and 2/5 patients had brachial plexus injuries or accompanying brachial plexus injuries after military training, push-ups, and backpack carriages, respectively. The electrodiagnostic patterns for each trigger are shown in Table 3.

**Table 3.** Triggering Activities and Electrodiagnostic Patterns.

| Electrodiagnostic Patterns<br>N (%) | Triggering Activities<br>N (%) | Military training | Push-ups | Backpack<br>carriages | Sleep | Trauma | Unknown |
| --- | --- | --- | --- | --- | --- | --- | --- |
|  |  | 16 (25.0) | 13 (20.3) | 7 (10.9) | 6 (9.4) | 4 (6.3) | 18 (28.1) |
| SMDPN<sup>□</sup> | 47 (73.4) | 9 | 8 | 5 | 4 | 3 | 18 |
| SMDPN + Brachial plexopathy | 8 (12.5) | 4 | 3 |  | 1 |  |  |
| Brachial plexopathy | 7 (10.9) | 2 | 2 | 2 | 1 |  |  |
| Mononeuropathy | 2 (3.1) | 1 |  |  |  | 1 |  |

□ SMDPN, Sensorimotor demyelinating polyneuropathy

We analyzed in detail the nerve conduction study results of 41 patients who underwent neurophysiological testing at our center. The tests were performed at a mean of 7.3 (1–44) months after symptom onset, and the results were compared with those of previous investigations of nerve conduction studies in patients with HNPP (Table 4).^2,3,10,11)^

**Table 4.**
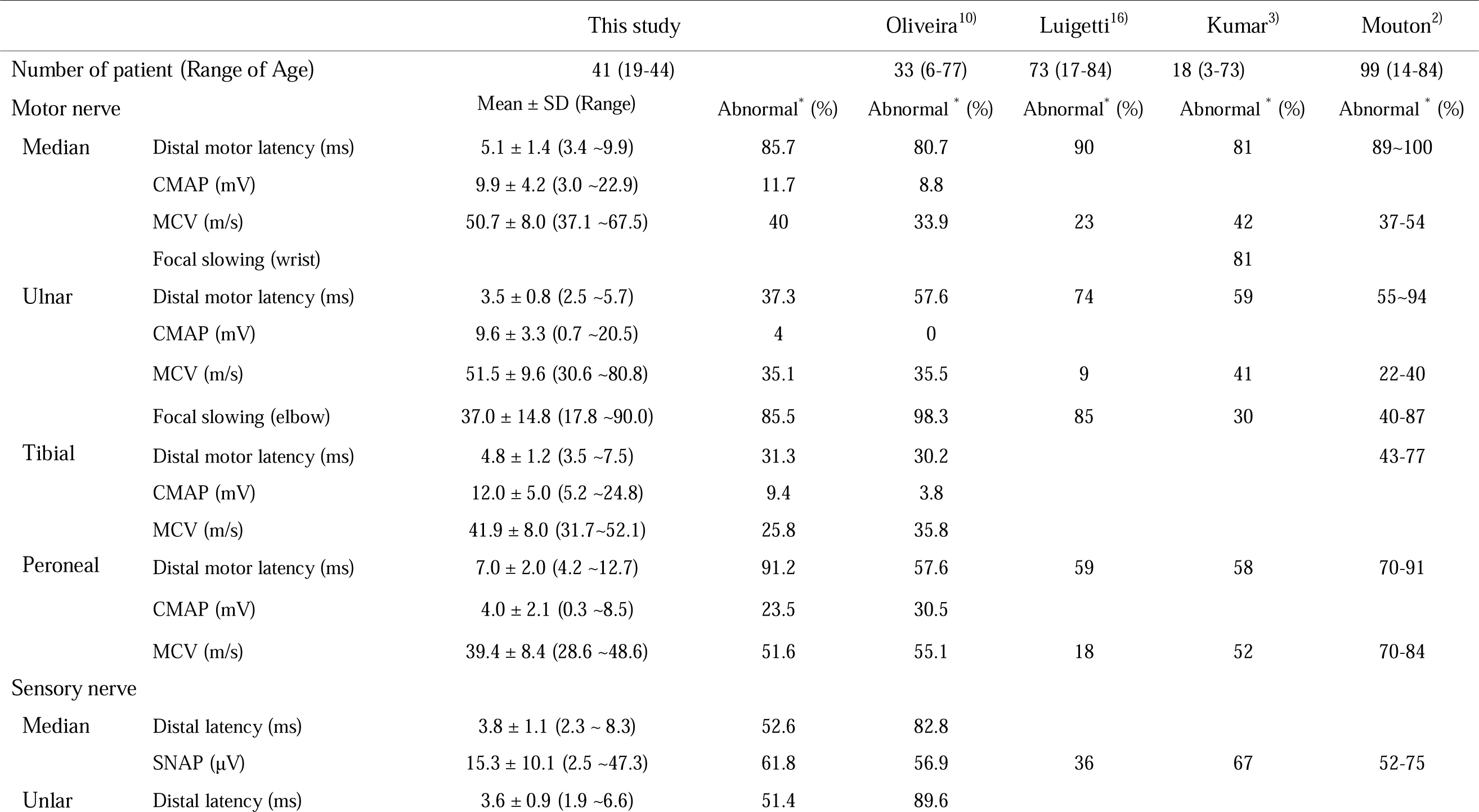
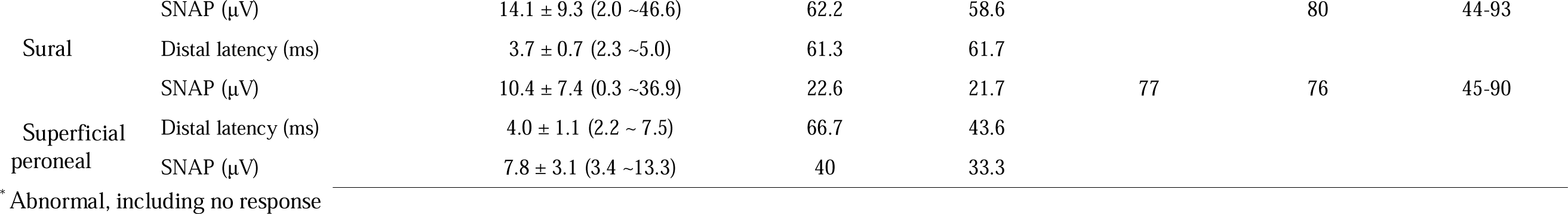
Results of the nerve conduction studies compared with previous studies.

Delayed distal motor latency or no response of the peroneal nerve was observed in 91.2% of patients with HNPP, and delayed distal motor latency or no response of the median nerve was observed in 85.7%. In addition, a motor nerve conduction study revealed focal slowing across the elbow in the segment or no response of the ulnar nerve in 85.5% of patients.

There was no statistically significant difference between the left and right motor and sensory nerve conduction studies (Tables 5 and 6). Abnormal spontaneous activity was observed in 27 of the 36 patients who underwent needle electromyography.

**Table 5.**
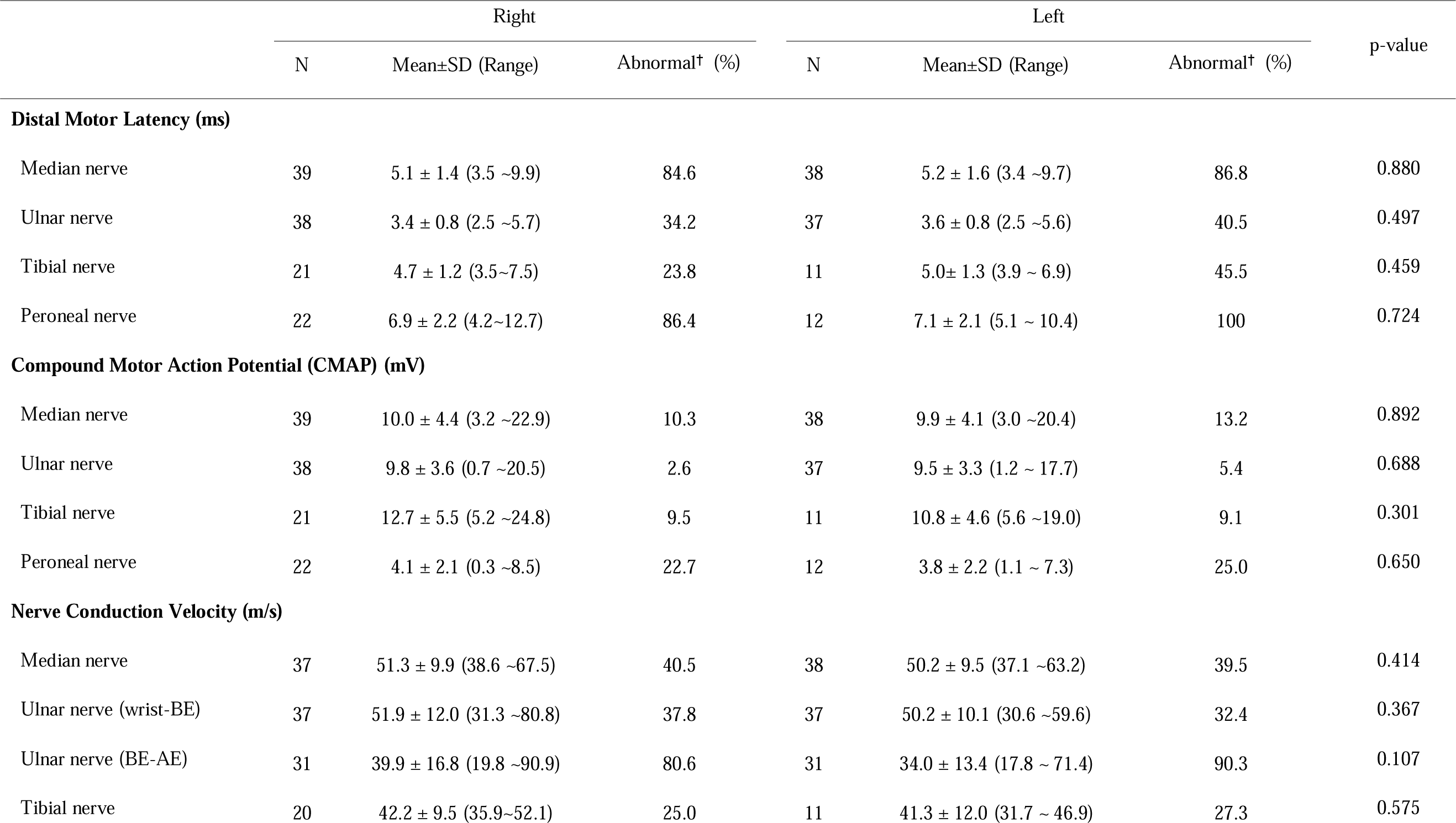
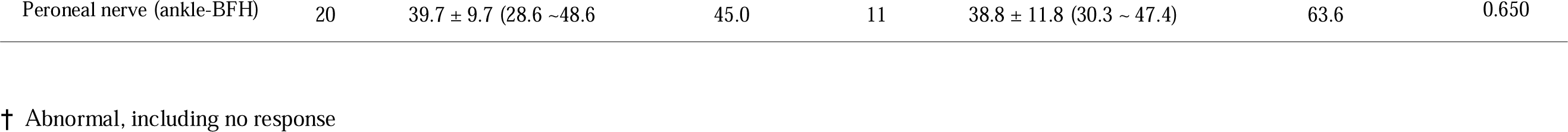
Results of the motor nerve conduction studies.

|  | Right |  |  | Left |  |  | p-value |
| --- | --- | --- | --- | --- | --- | --- | --- |
|  | N | Mean±SD (Range) | Abnormal† (%) | N | Mean±SD (Range) | Abnormal† (%) |  |
| Distal Motor Latency (ms) |  |  |  |  |  |  |  |
| Median nerve | 39 | 5.1 ± 1.4 (3.5 ~9.9) | 84.6 | 38 | 5.2 ± 1.6 (3.4 ~9.7) | 86.8 | 0.880 |
| Ulnar nerve | 38 | 3.4 ± 0.8 (2.5 ~5.7) | 34.2 | 37 | 3.6 ± 0.8 (2.5 ~5.6) | 40.5 | 0.497 |
| Tibial nerve | 21 | 4.7 ± 1.2 (3.5~7.5) | 23.8 | 11 | 5.0± 1.3 (3.9 ~ 6.9) | 45.5 | 0.459 |
| Peroneal nerve | 22 | 6.9 ± 2.2 (4.2~12.7) | 86.4 | 12 | 7.1 ± 2.1 (5.1 ~ 10.4) | 100 | 0.724 |
| Compound Motor Action Potential (CMAP) (mV) |  |  |  |  |  |  |  |
| Median nerve | 39 | 10.0 ± 4.4 (3.2 ~22.9) | 10.3 | 38 | 9.9 ± 4.1 (3.0 ~20.4) | 13.2 | 0.892 |
| Ulnar nerve | 38 | 9.8 ± 3.6 (0.7 ~20.5) | 2.6 | 37 | 9.5 ± 3.3 (1.2 ~ 17.7) | 5.4 | 0.688 |
| Tibial nerve | 21 | 12.7 ± 5.5 (5.2 ~24.8) | 9.5 | 11 | 10.8 ± 4.6 (5.6 ~19.0) | 9.1 | 0.301 |
| Peroneal nerve | 22 | 4.1 ± 2.1 (0.3 ~8.5) | 22.7 | 12 | 3.8 ± 2.2 (1.1 ~ 7.3) | 25.0 | 0.650 |
| Nerve Conduction Velocity (m/s) |  |  |  |  |  |  |  |
| Median nerve | 37 | 51.3 ± 9.9 (38.6 ~67.5) | 40.5 | 38 | 50.2 ± 9.5 (37.1 ~63.2) | 39.5 | 0.414 |
| Ulnar nerve (wrist-BE) | 37 | 51.9 ± 12.0 (31.3 ~80.8) | 37.8 | 37 | 50.2 ± 10.1 (30.6 ~59.6) | 32.4 | 0.367 |
| Ulnar nerve (BE-AE) | 31 | 39.9 ± 16.8 (19.8 ~90.9) | 80.6 | 31 | 34.0 ± 13.4 (17.8 ~ 71.4) | 90.3 | 0.107 |
| Tibial nerve | 20 | 42.2 ± 9.5 (35.9~52.1) | 25.0 | 11 | 41.3 ± 12.0 (31.7 ~ 46.9) | 27.3 | 0.575 |
| Peroneal nerve (ankle-BFH) | 20 | $39.7 \pm 9.7$ (28.6 ~48.6) | 45.0 | 11 | $38.8 \pm 11.8$ (30.3 ~ 47.4) | 63.6 | 0.650 |

† Abnormal, including no response

**Table 6.**
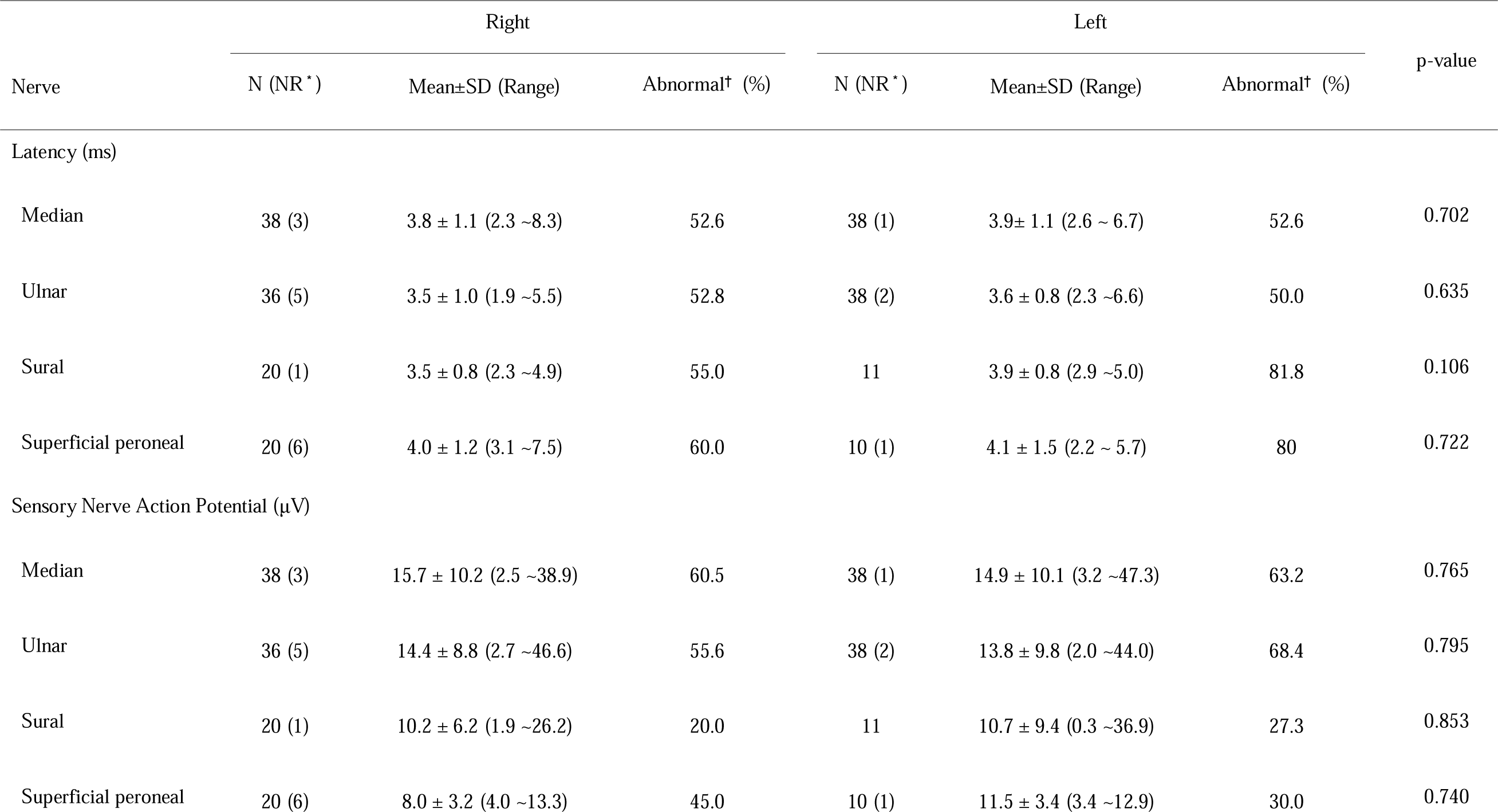
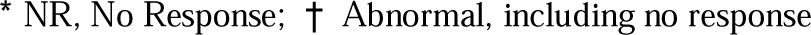
Results of the sensory nerve conduction studies.

| Nerve | Right |  |  | Left |  |  | p-value |
| --- | --- | --- | --- | --- | --- | --- | --- |
|  | N (NR<sup>+</sup>) | Mean±SD (Range) | Abnormal<sup>†</sup> (%) | N (NR<sup>+</sup>) | Mean±SD (Range) | Abnormal<sup>†</sup> (%) |  |
| Latency (ms) |  |  |  |  |  |  |  |
| Median | 38 (3) | 3.8 ± 1.1 (2.3 ~8.3) | 52.6 | 38 (1) | 3.9± 1.1 (2.6 ~ 6.7) | 52.6 | 0.702 |
| Ulnar | 36 (5) | 3.5 ± 1.0 (1.9 ~5.5) | 52.8 | 38 (2) | 3.6 ± 0.8 (2.3 ~6.6) | 50.0 | 0.635 |
| Sural | 20 (1) | 3.5 ± 0.8 (2.3 ~4.9) | 55.0 | 11 | 3.9 ± 0.8 (2.9 ~5.0) | 81.8 | 0.106 |
| Superficial peroneal | 20 (6) | 4.0 ± 1.2 (3.1 ~7.5) | 60.0 | 10 (1) | 4.1 ± 1.5 (2.2 ~ 5.7) | 80 | 0.722 |
| Sensory Nerve Action Potential (μV) |  |  |  |  |  |  |  |
| Median | 38 (3) | 15.7 ± 10.2 (2.5 ~38.9) | 60.5 | 38 (1) | 14.9 ± 10.1 (3.2 ~47.3) | 63.2 | 0.765 |
| Ulnar | 36 (5) | 14.4 ± 8.8 (2.7 ~46.6) | 55.6 | 38 (2) | 13.8 ± 9.8 (2.0 ~44.0) | 68.4 | 0.795 |
| Sural | 20 (1) | 10.2 ± 6.2 (1.9 ~26.2) | 20.0 | 11 | 10.7 ± 9.4 (0.3 ~36.9) | 27.3 | 0.853 |
| Superficial peroneal | 20 (6) | 8.0 ± 3.2 (4.0 ~13.3) | 45.0 | 10 (1) | 11.5 ± 3.4 (3.4 ~12.9) | 30.0 | 0.740 |

\* NR, No Response; † Abnormal, including no response

### Genetic study

As a results of test for the PMP22 gene, deletions were observed in 61 patients and mutations were observed in 3 patients. The mutations include a pathogenic variant (c.79-2A>C) in one patient at the exon 3 acceptor splice site, a splice site variant (c.179-1G>C) in one patient at the intron 3 site, and a splice site variant (c.78+1G>C) in one patient at the intron 2 site.

## DISCUSSION

HNPP often occurs during military service and it is more prevalent in soldiers than in the general population due to its predominance in males and the high level of physical activity. In a study of Korean, 31.3% of patients with HNPP reported symptom onset during military service.^9)^ A study involving Finnish military personnel reported brachial plexus palsy due to backpack carriage in 53.7 per 100,000 individuals, with HNPP diagnosed in approximately 7% of these cases.^4)^ Thus, the prevalence of HNPP affecting the brachial plexus in Finnish military personnel can be estimated to be at least 3.8 per 100,000. Another study on the prevalence of neuromuscular disorders in Korean men in their early 20s reported 78 cases of HNPP in the South Korean military over a 10-year period, resulting in a prevalence of 3.1 per 100,000.^12)^

Although previous studies have focused on brachial plexus injuries for HNPP occurring in soldiers, but 54.7% of patients were presenting with various forms of mononeuropathy and 28.1% presenting with brachial plexopathy in this study. Having a good understanding of the phenotype associated with the triggering activity of HNPP can aid clinicians in considering HNPP as a diagnosis in patients presenting with abnormal neurological symptoms based on their history. The most common triggering activity of mononeuropathy was military training. We found that if the onset occurred after a shoulder-intensive activity, such as push-ups and backpack carriages, the phenotype was often manifested in the form of brachial plexus injury.

In this study, most patients with HNPP had weakness due to a typical presentation of focal nerve palsy, but 17.2% of them exhibited an atypical clinical presentation.^2,11)^ Two of the three patients with a Guillain–Barre-like presentation, which is a relatively well-known atypical clinical presentation of HNPP, were misdiagnosed with Guillain–Barre syndrome and received immunotherapy, but the symptoms did not improve, thereby diagnosis of HNPP.

Although not reported in previous studies, we observed a lumbosacral radiculopathy-like presentation as an atypical clinical presentation of HNPP. In patients who complained of lower back pain and radiating leg pain, herniations of lumbar discs were observed in radiology tests, and they were diagnosed with lumbosacral radiculopathy. One of the three patients underwent lumbar fusion, but the symptoms did not improve and the patient was diagnosed with HNPP on further thorough examination.

A better understanding of the atypical clinical presentation of HNPP will also help in early diagnosis, as atypical presentations of HNPP can lead to delayed diagnosis or misdiagnosis as other conditions, potentially resulting in unnecessary surgery.

Previous studies and studies involving military populations have reported the most abnormalities in motor nerve conduction studies of the median nerve in HNPP.^1,2,3,10,11)^ However, when we analyzed the nerve conduction studies of 41 patients with HNPP who underwent electrophysiologic studies, it was found that the peroneal motor nerve had the highest incidence of abnormalities including no response, with 91.2% of distal latency, in contrast to previous studies.^,3,10,16)^ The reason for most patients complaining of upper extremity symptoms but exhibiting more abnormalities in the distal latency of the peroneal motor nerve can be attributed to the fact that soldiers spend significant time walking due to marching or training, which often stimulates the lower extremity nerves, and that soldiers wear combat boots that can compress the nerves in the lower leg.

In addition, axonal involvements were frequently observed, with abnormal spontaneous activity observed in 75% of patients who underwent needle electromyography. This may be because in HNPP patients with genetically vulnerable nerves, minor trauma during physical activities such as military training or drills can lead to rapid demyelination or focal axonal damage.

Based on the findings of this study, there are several considerations when clinically suspecting HNPP in soldiers and performing electrophysiologic study. First, even if the patient complains of symptoms in the upper extremities, the inclusion of motor nerve conduction studies of the peroneal nerve may help in the early diagnosis of HNPP. Second, it is important to consider that in physically active soldiers, HNPP is often accompanied by focal axonal damage.

The results of this study showed that special activities play significant a role in the manifestation of symptoms in soldiers with HNPP, and that different nerves are affected depending on the triggering activity, leading to a variety of phenotypes.

However, there are some limitations to this study. First, the retrospective nature of the study and the reliance on medical records may introduce biases and limit the generalizability of our findings. Second, since the subject of the study is a soldier in a particular country, the results of this study may not be generalized and applied to otr soldiers who use different equipment and receive different physical training. Lastly, since the diagnostic criteria was limited to cases confirmed by genetic testing, this study may present different clinical features and electrodiagnostic patterns compared to previous studies.

## Contributorship Statement

The author solely conceived, designed, collected data, analyzed, and wrote the manuscript. The author is the guarantor of this work and takes full responsibility for the overall content and the integrity of the data presented.

## Data Availability

All data produced in the present work are contained in the manuscript

